# Rapid Response for Notification of Monkeypox Exposure, Exposure Risk Assessment and Stratification, and Symptom Monitoring

**DOI:** 10.1101/2022.06.12.22275974

**Authors:** Lynn A. Simpson, Kaitlin Macdonald, Eileen F. Searle, Jennifer A. Shearer, Dimitar Dimitrov, Daniel Foley, Eduardo Morales, Erica S. Shenoy

## Abstract

**Objective:** Public health authorities recommend symptom monitoring of healthcare personnel (HCP) after defined exposures to Monkeypox (MPX). We report on the rapid development and implementation of mobile responsive survey solutions for notification of possible exposure, exposure risk assessment and stratification, and symptom monitoring.

**Setting:** Academic health center in Boston, Massachusetts, after admission of first diagnosed case of MPX in the United States during the current global outbreak.

**Participants:** Research Electronic Data Capture (REDCap) design and programmers, Infection Control, Occupational Health, and Emergency Preparedness specialists, and HCP with possible exposure to MPX.

**Interventions:** Design and deployment of Research Electronic Data Capture (REDCap) tools to identify HCP with possible exposure to MPX, perform exposure risk assessment and stratification for post-exposure prophylaxis (PEP), and conduct symptom monitoring during the exposure window. Project enhancements included dashboards for HCP tracking and SMS text reminders for symptom monitoring.

**Results:** Tools to support the contact tracing and exposure investigation were deployed within 24 hours of identification of a patient with suspected MPX, with the full suite in production within 4 days of confirmation of the diagnosis of MPX. Clinical follow up of HCP was integrated into the design, and real-time versioning allowed for adjustments leading to improvements in HCP symptom monitoring compliance and enhanced tracking.

**Conclusions:** During the current MPX outbreak, timely and comprehensive evaluation of potential HCP exposures is necessary but presents logistical challenges. Rapid development of MPX-specific solutions using REDCap enabled flexibility in design and approach, and integration of targeted clinical support enhanced functionality.

## INTRODUCTION

As the outbreak of Monkeypox (MPX) outside of endemic areas continues,^1^ healthcare facilities and public health authorities face the challenge of rapid identification of individuals with possible exposures to MPX and subsequent management. While there are various definitions of exposure developed by the United Kingdom,^2^ Centers for Disease Control and Prevention (CDC),^3^ and World Health Organization,^4^ the challenges of efficient notification of possible exposure, performance of a risk assessment to allow for risk stratification, and symptom monitoring during the exposure window are substantial. Of immediate importance is risk stratification because individuals at higher risk may be offered post-exposure prophylaxis (PEP) in the form of vaccination, however the optimal window to offer vaccination is within 4 days of exposure.^5^

We developed and deployed a suite of tools using REDCap (Research Electronic Data Capture) as part of our MPX response building on prior use supporting planning and preparedness efforts at Massachusetts General Hospital (MGH) and within the Mass General Brigham (MGB) system. The REDCap technology was in use prior to the current MPX outbreak as part of planned monitoring of HCP caring for patients in our facilities’ Biocontainment Unit and the Special Pathogens Unit at the Region 1 Emerging Special Pathogens Treatment Center. Use of this technology was further developed to support MGB during the COVID-19 pandemic, including use for symptom attestation prior to on-site work for HCP,^6^ symptom monitoring after COVID-19 vaccination,^7-9^ and creation of survey tools for COVID-19 exposure notification, risk assessment and stratification. REDCap is a secure, web-based software platform designed to support data capture for research studies.^10,11^

We report on the rapid development of MPX-specific solutions and early experience in deploying the technology including 1) Notification of Possible Exposure; 2) Exposure Risk Assessment and Stratification, and 3) Symptom Check, as well as access to these tools for use by others. Each solution was additionally enhanced by clinical support teams providing HCP with counseling and guidance.

## METHODS

Three tools were developed: 1) Notification of Possible Exposure; 2) Exposure Risk Assessment and Stratification, and 3) Symptom Check. The flow of data into and between these tools is depicted (**Figure 1**).

**Figure 1.**
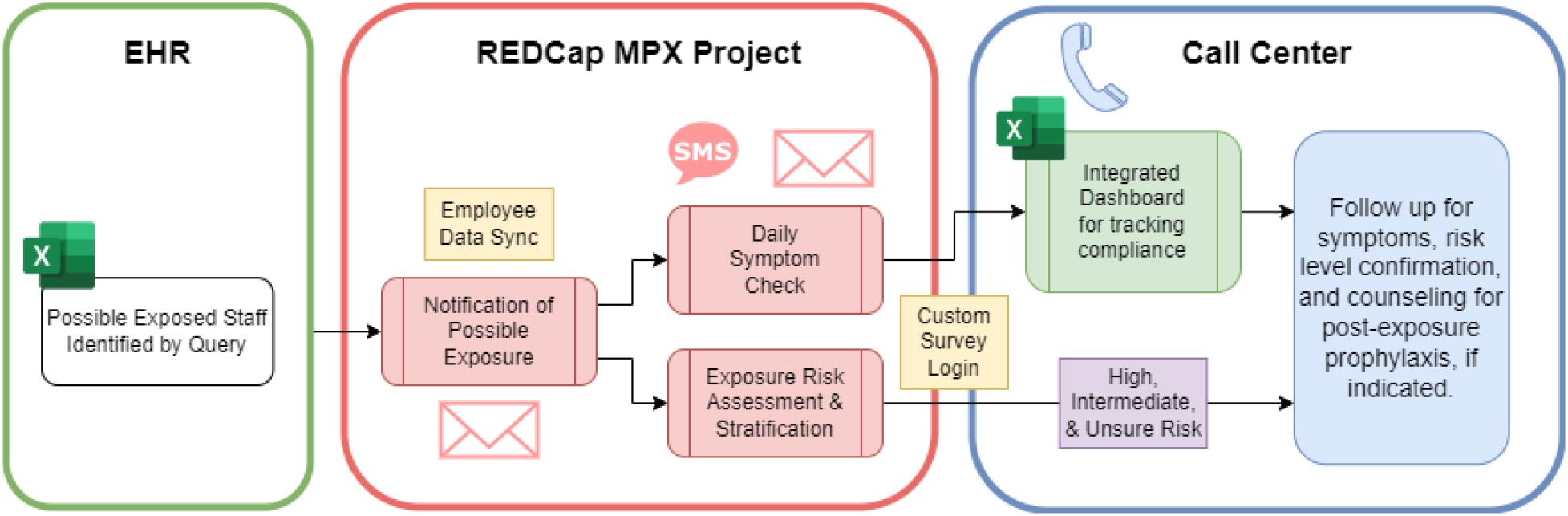
Data Architecture of MGH Monkeypox REDCap Toolkit. Contact trace data from the Electronic Health Record (EHR) was extracted from MS Excel and imported into REDCap. HCP received the Notification of Possible Exposure Survey, and if they attested “yes” they proceeded to Symptom Check and Exposure Risk Assessment and Stratification surveys. Call Center staff utilized the Integrated Dashboard to track compliance and establish follow up for symptoms, as well as risk level confirmation, and counseling for post-exposure prophylaxis, if indicated. The REDCap External Modules (Employee Data Sync and Custom Survey Login) were used to enhance the project functionality.

### Determination of the At-Risk Population

HCP were initially identified as possibly exposed through pre-established standard processes used for contact tracing and exposure investigations at our institution, utilizing a trace within the electronic health record (EHR), and supplemented with interviews with managers conducted by Infection Preventionists (IPs) and Occupational Health Services (OHS) clinicians. The dataset included first and last exposure dates and was imported into a REDCap project and combined with additional HCP data using a custom MGB REDCap external module developed previously to support OHS operations. Additional HCP data included contact information such as email address and phone numbers.

### REDCap Build

The tools were developed using all native REDCap v12.0.19 functionality except for two MGB custom External Modules: Employee Data Sync and Custom Survey Login. External Modules are individual software packages that are created and installed by a REDCap developer/administrator. REDCap provided technical support for the core project needs; however, due to the need for rapid assessment to deploy PEP, and complexity of exposure assessment, the establishment of a team of clinicians to support HCP use of all three tools was determined to be necessary and a Call Center was established. The Call Center staff were identified and trained via a Labor Pool that was established as part of MGH’s Hospital Incident Command System which was mobilized to support this response. External Modules extended current functionality of REDCap and, for this project, enhanced data workflow, and allowed Call Center staff to login and initiate a record on behalf of the HCP.

The REDCap build consisted of multiple surveys, email and SMS notifications, and an advanced approach combining custom logic utilizing @CALCTEXT action-tag with nested IF-statements for providing dynamic feedback to inform HCP of the results of their Exposure Risk Assessment and Stratification and Symptom Check. For Symptom Check compliance, the Call Center utilized an MS Excel file that was programmatically updated daily with HCP data exported from REDCap via an Application Programmable Interface (API).

This project was conducted as part of Infection Control and Occupational Health Services and not subject to human subjects review.

## RESULTS

Three integrated modules were developed within REDCap. Detailed specifications of the tools, including an implementation guide is provided (Supplement), including instructions on accessing the project at REDCap and github.

### Notification of Possible Exposure Tool

In order to both identify HCP who could have been exposed to the index patient, and to exclude HCP with no possible exposure, all HCP on the comprehensive trace list received the Notification of Possible Exposure Survey tool.

The trace list included first and last exposure dates and was exported from the EHR to an MS Excel file and then imported into REDCap using the Data Import tool. These data were combined with additional HCP data, email address and phone numbers, using the custom MGB REDCap external module: Employee Data Sync. Once an email address was available, REDCap automatically sent an email to the HCP with information about the type of exposure, dates, and locations and allowed the HCP to attest to preliminary exposure question (yes/no). If the HCP indicated “Yes” to the preliminary question, they were enrolled into a symptom monitoring tool: Symptom Check. This enrollment occurred prior to exposure risk assessment and stratification. Within approximately 24 hours of preliminary diagnosis of the patient with MPX, the Notification of Possible Exposure surveys were created and emailed to the first set of identified HCP and HCP began to respond immediately.

### Exposure Risk Assessment and Stratification Tool

Once HCP had been identified as meeting a preliminary definition of MPX exposure based on response to the Notification of Possible Exposure, the next step was risk assessment and classification. Applying a framework based on the CDC^3^ and developed in collaboration with the Massachusetts Department of Public Health, the risk stratification was finalized on May 20, 2022, and integrated into the REDCap design over the next day and a half to complete the build on May 22, 2022. HCP were presented with a series of exposure scenarios and asked to identify which one(s) applied to their interactions with the index patient. Based on their responses, each HCP was categorized as High, Intermediate, or Low/Uncertain Risk. Individuals who answered “No” to all were recategorized as No Risk and were removed from the investigation and from Symptom Check. Individuals who reported being unsure were contacted by the Call Center staff to review their case and determine their risk classification.

The survey displayed each category’s questions as a progression. Starting with the High Risk category questions, if HCP indicated yes/unsure, they did not proceed to the next category and questions did not display for Intermediate or Low/Uncertain Risk. If HCP indicated no for the High Risk category, they proceeded to Intermediate and so on. Multiple hidden fields on the survey were used to calculate the risk category and number of days from last exposure (I.e., the exposure window). These derived variables were used to display specific text to the HCP utilizing @CALCTEXT action-tag with nested IF-statements (I.e., if PEP was recommended or not).

### Symptom Check Tool

Individuals identified in High, Intermediate, and Low/Uncertain Risk classifications required symptom monitoring for 21 days from their last exposure per public health guidance. While HCP at our healthcare facility were required to attest to be asymptomatic with respect to COVID-19 symptoms prior to on-site work,^6^ the symptoms from MPX included those not part of our standard COVID-19 symptom screening, and the immediate actions based on a positive symptom screen differed.

The Symptom Check survey included information about symptom monitoring for Monkeypox, the HCP symptom monitoring start and end date, and asked “Since your last survey, have you had the following symptoms”. Symptom questions included fever, chills, new lymphadenopathy and new skin rash. If the HCP answered yes to any symptom, the survey would display instructions to self-isolate and contact OHS immediately. This information was also automatically emailed to HCP. Reports were available in REDCap to OHS and the Call Center staff to immediately identify any HCP reporting symptoms. Symptom Check compliance was encouraged by multiple methods: email, calls and text messaging.

Compliance with symptom monitoring was assessed: 67.0% of HCP were compliant with their Symptom Check 100% of surveyed days. Among the remaining HCP, Symptom Check was missed, on average, for 4.1 days during the exposure window to date; these HCP received follow up calls from Call Center and OHS staff. Employees Symptom Check start dates varied and many last exposure dates were before the 21 day window (i.e., before implementation of the survey). Therefore, not all employees were required to complete the full 21 days and some employees with re-exposures were required to complete more than 21 days (i.e., those who continued to provide care to the patient and were in the Low/Uncertain category).

### Integrating Clinical Follow Up Into Design

REDCap provided technical support for the core project needs, however, due to the need for rapid assessment to deploy PEP, and complexity of exposure assessment, establishment of a Call Center was needed. This workflow addressed HCP with limited technical proficiency, language barriers, or both, as well as focused review of HCP reporting High or Intermediate Risk exposures who may be offered PEP.

### Timeline of Implementation and Versioning in Real-Time

The initial identification of a possible case of MPX occurred on May 17, 2022, with confirmation by the CDC on May 18, 2022. The first Notification of Possible Exposure was sent at 8:21pm on May 18, 2022, and initial Symptom Checks began at 5:54pm on May 19, 2022. Upon confirmation of risk exposure categorization and stratification with Massachusetts Department of Public Health on May 20, 2022, the first Exposure Risk Assessments and Risk Stratifications were deployed via phone call on May 22, 2022. (Figure 2)

**Figure 2.**
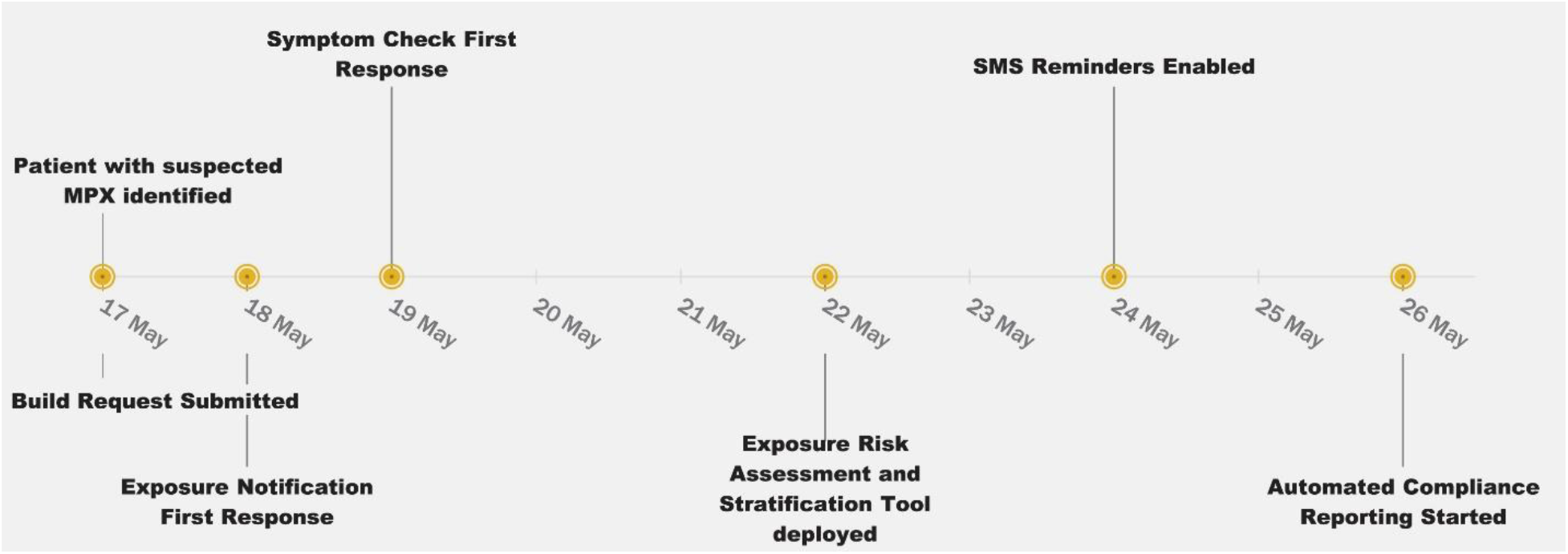
Timeline of Implementation for MGH Monkeypox REDCap Toolkit. Within hours of identification of a suspect MPX case at Massachusetts General Hospital, a request for REDCap build was submitted. The Notification of Possible Exposure tool was deployed within 24 hours of preliminary diagnosis of MPX, and the Symptom Check Tool shortly thereafter, with first responses received within 24 hours of confirmation of MPX diagnosis. The Exposure Risk Assessment and Stratification Tool was deployed on May 22, 2022 followed by SMS Symptom Check reminders on May 24 and automated compliance reporting on May 26, 2022.

Versioning of assessments were included as part of the strategy to maintain data integrity and reporting consistency. The Symptom Check was reduced from twice daily to once daily during the study period and additional functionality was added to support HCP including SMS (text message) reminders and by expanding the call center’s scope. Adding in the SMS option for daily reminders resulted in an increase in compliance. HCP who enrolled in SMS messaging had an average daily compliance rate of 94.9%, while those only receiving email reminders had a compliance rate of 88.4%.

### Support for HCP with limited technical proficiency or language barriers

For each tool, managers assisted in identifying a subset of HCP who, for either technical or language barriers, or both, required one-on-one assistance with the tools. Call Center clinicians contacted HCP by phone to provide information about the MPX Exposure Risk Assessment and completed the HCP assessment.

### Support for HCP with Reported High and Intermediate Risk Exposures

HCP reporting High and Intermediate Risk exposures received a prompt upon survey completion indicating that they would receive a follow up by Call Center clinicians to assist in exposure verification and categorization, and to provide PEP counseling, when indicated.

### Integrated Dashboard for Tracking

For Symptom Check compliance, the Call Center utilized an MS Excel file that was programmatically updated daily with HCP data exported from REDCap via an Application Programmable Interface (API). Survey data collected in REDCap was loaded into an MS Excel file via a Power Query accessing the REDCap API. A transformation of the data was preprogrammed that would produce a final dataset of non-compliant HCP and their associated contact information. Call Center staff were trained on how to easily perform the extract, which required no knowledge of coding, advanced excel knowledge, or authentication into REDCap. Call Center staff were trained on how to document if a phone call was made and any notes that may assist in future efforts.

Additionally, many reports were created within REDCap that allowed stakeholders and technical staff the ability to monitor the actual symptom data that was being generated. Most notably a report was created that would return any HCP who had reported being symptomatic at their latest assessment.

## DISCUSSION

Timely and comprehensive evaluation of exposures, including risk assessment, and ongoing symptom monitoring is necessary and can present logistical challenges, especially in the setting of an emerging outbreak with evolving definitions of exposures and urgency to offer and deploy PEP for a subset of exposed individuals. We report on rapid development of MPX-specific solutions using REDCap and early experience in deployment.

Utilizing REDCap as the base technology enabled flexibility in design and approach, and integration of targeted clinical support enhanced the functionality of the tools developed. The technology used by the clinical team supporting HCP had to be intuitive, accessible in real time by multiple remote staff, provide search abilities for HCP records, and a data entry interface. Using another previously developed custom MGB REDCap external module, assigned team members were able to access a user-friendly interface to search for HCP records within the REDCap project. Proactive identification of HCP who may benefit from clinical support, either based on technical proficiency (including both access to email and comfort with survey completion) or language barriers, or for whom counseling for PEP is indicated, was a critical component of the approach. As the tools continue to be deployed, workflows including clinical support can continue to be optimized for efficiency.

The MGH Monkeypox REDCap Toolkit and any custom external modules code are available to other organizations to use (Supplement). Customization may be indicated if an organization or entity is adapting local public health definitions of exposure or follow up based on responses. The overall strategy of the solution design and development was collaborative, with different developers creating components of the solution simultaneously by focusing on specific areas: survey question formats, automated notifications, authorized access and reporting.

In summary, there was an immediate need to operationalize exposure notification, risk assessment and stratification, and symptom monitoring technology within days of identification of a case of MPX. This was possible due to the modularized nature of REDCap, the concise communications from the teams involved including OHS and Infection Control on their system needs, and the experience of the development team in creating similar tools prior to and during the COVID-19 pandemic to support operations and research efforts. We anticipate that sharing this approach and the tools employed may assist others in similar circumstances, and lead to advancements and improvements. There is also broad applicability of these tools outside of healthcare settings, where many of the same needs have been identified in the response to community spread of MPX.

## Data Availability

All data produced in the present work are contained in the manuscript

https://community.projectredcap.org/articles/128815/mgh-monkeypox-redcap-toolkit.html

## ACKNOWLEDGEMENTS

The authors would like to thank the members of the MGH MPX Response Team, including healthcare personnel from Patient Care Services, Occupational Health Services, Infection Control, Emergency Preparedness, and the MGB REDCap Team.

