## Supplement for "Rapid Response for Notification of Monkeypox Exposure, Exposure Risk Assessment and Stratification, and Symptom Monitoring"

**Running title:** Rapid Response for MPX: REDCap to the Rescue

**Authors**
Lynn A. Simpson, MPH[1,2]
Kaitlin Macdonald, MSN, ANP-BC, COHN-S[3]
Eileen F. Searle, PhD, RN, CCRN [4,6]
Jennifer A. Shearer, MPH [4,5]
Dimitar Dimitrov, MSSE [2]
Daniel Foley, BA [2]
Eduardo Morales, PhD [2]
Erica S. Shenoy, MD, PhD*‡[6,7,8,9]

[1]Harvard Catalyst, The Harvard Clinical and Translational Science Center
[2]Mass General Brigham Research Information Science & Computing
[3]Mass General Brigham Occupational Health Services
[4]Center for Disaster Medicine, Massachusetts General Hospital
[5]Department of Emergency Preparedness and Business Continuity, Mass General Brigham
[6] Regional Emerging Special Pathogens Treatment Center, Massachusetts General Hospital
[7] Infection Control Unit, Massachusetts General Hospital, Boston, MA
[8] Division of Infectious Diseases, Department of Medicine, Massachusetts General Hospital
[9] Harvard Medical School

All in Boston, MA

**SUPPLEMENT**

**‡ Corresponding author:**
Erica S. Shenoy, MD, PhD
Massachusetts General Hospital
Infection Control Unit
55 Fruit Street, Bulfinch 334
Boston, MA, 02114


**About REDCap**
REDCap (**R**esearch **E**lectronic **D**ata **Cap**ture), https://www.project-redcap.org/ Vanderbilt University, with collaboration from a consortium of academic and non-profit institutional partners, develops this software application for electronic collection and management of research and clinical study data.[1,2] For non-profit and not for profit organizations, the licensing is free. More info is available to REDCap administrators: https://projectredcap.org/partners/termsofuse/ Over 5900 institutions and 145 countries have implemented REDCap at their institutions.

**Accessing the MGH Monkeypox REDCap Toolkit**
The MGH Monkeypox REDCap Toolkit will be available to the REDCap Community. The build will be shared via

1. the REDCap Consortium site at https://community.projectredcap.org/articles/128815/mgh-monkeypox-redcap-toolkit.html. Note that this location is only available to licensed institutions' REDCap administrators.
2. We have also posted materials on github at https://gitlab-scm.partners.org/mgb-redcap/mgh-mpx-redcap-toolkit.

The entire project (instruments, fields, and project attributes) can be downloaded as a single XML file (CDISC ODM format). This XML file can be used to create a clone of the project (no data) on another REDCap server (it can be uploaded on the Create New Project page). Because it is in CDISC ODM format, it can also be used to import the project into another ODM-compatible system.

**MGH MPX REDCap Project Build**
This supplement explains in more detail the REDCap project design and build.

*Project Instruments*
An Employee Info instrument contained data collection fields for healthcare personnel (HCP) contact information and associated metadata needed for triggering or cancelling alerts. This form was also utilized for the Custom Survey Login external module landing page. Participant specific notes were also kept in this instrument.

The Notification of Possible Exposure survey collected a preliminary exposure question. If the HCP indicated “Yes” to the preliminary question, they were enrolled into the symptom monitoring tool, Symptom Check, using conditional logic.

The Exposure Risk Assessment and Stratification survey collected data on the level of exposure an HCP may have encountered. This instrument used a conditional approach to explore additional exposure risks depending on the items selected by the participant or screening staff. Branching logic allowed for conditional questioning, and @CALCTEXT / Calculated fields allowed for dynamic messaging and risk stratification. The survey queue was utilized to only display this instrument for completion if indicated by monitoring staff.

Daily symptom reporting was achieved by using the Symptom Check data collection instrument. This survey collected data on the current system status, as well as producing dynamic messaging and symptomatic status flags. This instrument was made a repeatable instrument, which would allow for a method to continually collect data over a 21-day period, but also permit extension of monitoring if required. The survey queue was utilized to prevent users from submitting repeating instances less than 4 hours apart.

*SMS Enrollment/SMS Opt-out instruments*

These were created to allow participants to select their messaging preference. Custom coded opt-in selection permits further functionality by displaying custom messaging in the survey queue and allowing confirmation of contact information. The survey queue would display opt-in options if they had not been selected, and opt-out

*MGB Custom External Module: Employee Data Sync.*

The MGH MPX data collection started with an EHR Query that returned a pre-defined list of HCP in MS Excel but did not include their email, phone number, or other required information. For Occupational Health staff, it is important to quickly and easily import the EHR Query data into REDCap and join it to additional HPC information. The External Module Employee Data Sync allows a REDCap project user to trigger a batch data sync to update all records or when entering individual data, allows data to sync on saving a record. The module is configured at the project level to 1) identify the field for the HCP unique Mass General Brigham (MGB) username and then 2) map additional fields from two external sources: Active Directory (LDAP) and Occupational Health and Human Resources Database (PeopleSoft, a human resources software in use at MGB). When a batch data sync is triggered or a record is saved, the module retrieves the HCP data from the sources, maps it to the designated project fields, and then updates the record(s). This module allowed the MGH MPX Response Team to quickly import potentially exposed HCP information in batches into REDCap with only the limited data set from the EHR Query, programmatically add email addresses to the records and immediately email survey notifications.

Alternatives to this External Module functionality include the ability to query an EHR for all the required HCP exposure and contact data to import into REDCap or if data is only available in multiple different sources, data can be imported in batches using MS Excel csv files.

*MGB Custom External Modules: Custom Survey Login*

By design, REDCap survey data collection is primarily dependent on sending email notifications with a participant specific survey link, providing navigation to a web page, or allowing for data entry. It was determined that a Call Center would be established to collect Exposure Risk Assessment and Stratification by phone for a subset of employees and to confirm risk stratification among HCP reporting High or Intermediate Risk exposures after completing the survey on their own. Call center staff are not routine users of REDCap, so a method was devised to allow them to enter data on behalf of other staff members, without needing to be familiar with the application.

MGB REDCap supports LDAP integration with Active Directory, however, this type of user authentication is not available at the survey-level. The External Module Custom Survey Login leverages the REDCap Public Survey link by adding an authentication screen linked to LDAP and providing a “Manager Portal”. A “Manager Portal” allows an authorized individual to login, search for an HCP by name, HCP ID or username, find and display the HCP MGH MPX REDCap record and survey queue, and to initiate survey data entry. The Call Center clinical staff was provided with a 15 minute training session on the use of the REDCap tools, and additional training on MPX, and a How To document and successfully accessed and executed the process within the hour and repeatedly throughout the Call Center implementation.

Alternatives to this functionality could not be determined by our team and therefore, it was coded as an External Module. The Custom Survey Login external module is widely used within MGB and is under active development. A version of the module is being prepared for release to the REDCap Consortium Repository. Additionally, other external modules with a similar functionality are available on the REDCap Repo.

*Dynamic Text for REDCap Survey Completion*

An advanced REDCap approach was used for providing dynamic feedback to inform HCP of next steps for all three MGH MPX tools. The ability to produce custom dynamic text can be leveraged by utilizing a REDCap feature referred to as “piping.” Piping allows data that has been either manually or produced programmatically to be presented in places such as survey completion pages, email/SMS alerts, and follow up questions.

During implementation, two approaches were identified for adding conditional logic to be presented to a participant. Either using a REDCap External Module (EM) that allows for custom JavaScript functionality to REDCap instruments or using the @CALCTEXT action-tag. The former, a procedural approach, is heavily dependent on JavaScript coding abilities and high-level administrative rights and permissions within REDCap. The latter, a conditional approach, requires deployment of data collection fields with @CALCTEXT action tags, available to all end users as of REDCap v10, which also require a lesser level of coding familiarity. These two methods proved to be equivalent, while the former was used for rapid prototyping and initial deployment, the latter was preferred as it minimizes resources for its long-term maintenance.

**Example of @CALCTEXT for Symptom Tracker**

```
@CALCTEXT(
if(
  [fever] = '2'
  or [chills] = '1'
  or [lymphadenopathy] = '1'
  or [rash] = '1',
  '<p><b>Action Required:</b> You have reported at least one symptom
that could be consistent with monkeypox. <p><b>Please self isolate
and contact Occupational Health Services immediately</b></p>',
  if(
    [fever] = '0'
```

```
        and [chills] = '0'
        and [lymphadenopathy] = '0'
        and [rash] = '0',
        '<p>You have reported no symptoms. <br><br> If you have any
    questions, please contact Occupational Health Services</p>',
        ''))) 
```

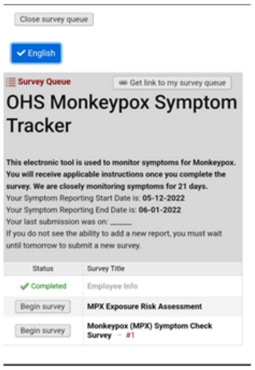


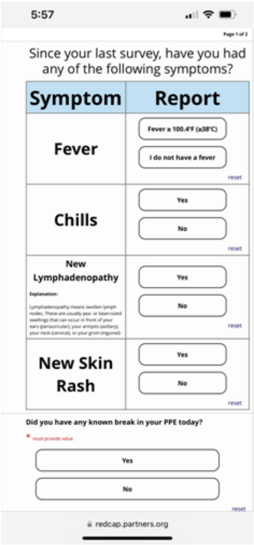


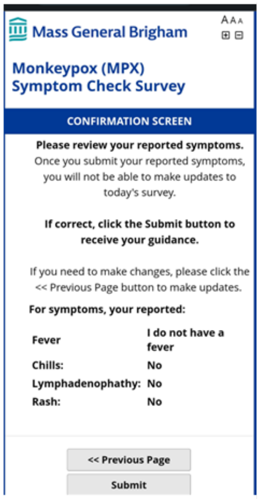


*Email Notifications: Alerts and Notifications Scheduling with Adjustable Start Date*

REDCap has multiple email scheduling capabilities and for the Symptom Check survey, we utilized Alerts & Notifications (Alerts) over Automatic Survey Invites (ASI). In addition to the initial email schedule of two times per day (AM and PM) for 21 days, the schedule was required to support potential re-exposure scenarios in which the HCP 21-day follow-up must restart from the date of latest exposure without losing previously collected data. Because of the re-exposure scenario, or more commonly when an HCP continued to care for a patient with MPX without known breaches in PPE and was considered in the Low/Uncertain Risk category, instead of a longitudinal project build, we opted to make the Symptom Check a repeating survey. The Alerts included a link to the Survey Queue so HCP could add a new survey when prompted.

```
Symptom Check AM Daily Reminder Alert Conditions
Step 1
A) When conditional logic is true during data entry
B) datediff([last_exposed],"today","d",true) <= "21" and
   [employee_info_complete] = 2 and [disable_am_alert] <>
   "1"
     a. Ensure Logic is still true before sending
        notification
C) Once Per Record
Step 2
```

- `Send Alert 1 minute after [am_alert_time]`
- `Send Every 1 days after initially being sent`
- `Send up to 50 times`

For REDCap v12.0.19, it is not possible to configure one alert that meets all these requirements, and through iterative design and testing, we determined multiple separate Alerts were necessary. Two separate Alerts were configured for AM and PM data collection. They were scheduled to be sent every day for 50 days to compensate for the potential re-exposure scenario. The alert was scheduled to be sent based on a calculated date-time field from the record. Conditional logic, including the expected positive logic, a stop-condition field, and a difference check between the current date and the last exposure date of the HCP ensured staff could be un-enrolled or extended. Conditional logic is evaluated before each alert is sent to prevent unwanted alerts.

The advantages of this design are that it allowed the HCP-specific record data to control the Alerts functionality, which lead to Alerts automatically adjusting to the HCP latest/updated data. Erroneous Alerts could be sent by REDCap if Step 2 above was set to a fixed start date. In addition, including the HCP last exposure date as a field in the Alerts conditional logic enabled the Alert to adjust its schedule until the allotted symptom reporting window had closed.

To simplify the complexity of the Alert configurations, and for future builds, we will create an initial Alert to send as of “now” and only once. This will also allow for custom “welcome” text for the first notification. All subsequent notifications will utilize the methodology above and option to start alerts on the “next day”.

*SMS (text) Notifications*

To encourage participation for the Symptom Check, an option for reminders via SMS messaging was offered via REDCap and the Twilio online service (REDCap+Twilio). This approach provided a cost-effective method to increase participant engagement and increase overall compliance with symptom reporting requirements.

The REDCap platform provides a convenient and low-cost method to engage participants via SMS messaging via the Twilio platform. The REDCap+Twilio service offers SMS messaging for less than one cent per message. The technical skills needed to activate and use the Twilio service are low, and the REDCap platform provides step by step instructions on retrieving the required information from a Twilio account and how to implement the required data collection fields to allow users to select if they would like to enroll in the service.

The associated phone number purchased was in the Boston area code to reduce impression of spam messaging by the HCP. To remain HIPAA compliant, Twilio account configurations were modified to prevent the association of phone numbers and messaging content. Configuration of settings provided by Twilio were applied to the REDCap project.

After implementation of the Twilio service, an SMS enrollment and subsequent unenrollment form was created within the project and displayed on the HCP Survey Queue. HCP were prompted to choose to receive daily alert reminders via email or both Email and SMS messaging. HCP who chose to opt-in to receive SMS reminders were sent an initial enrollment SMS message as well as daily SMS messages with a link to complete their Symptom Check surveys. HCP could stop SMS messages at any point by either completing an “Unenrollment” survey or by texting STOP to the messaging system.

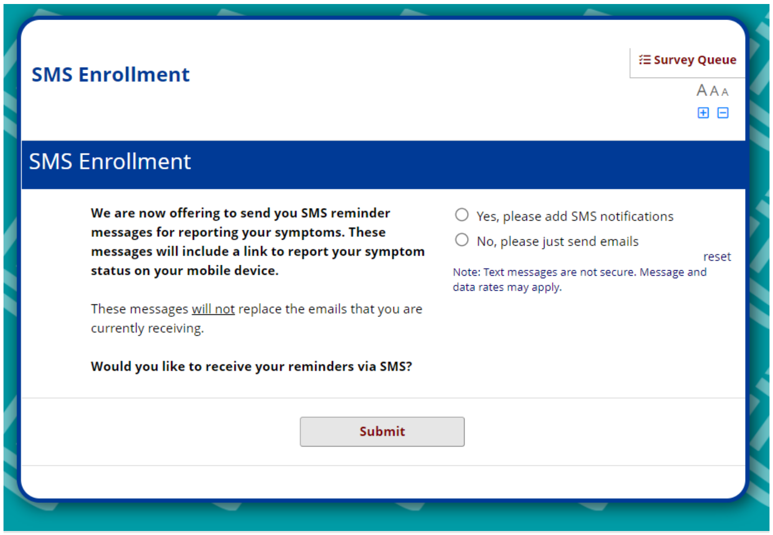


**REDCap SMS opt-in question values:**

| Raw Value | Label |
|---|---|
| SMS_INVITE_WEB | Yes, please add SMS notifications |
| EMAIL | No, please just send emails |

In addition to the email notifications outline above, two SMS based Alerts were created. The first was an SMS notification of enrollment and instructions on how to stop the SMS alerts. The second was a daily AM reminder SMS message with a hyperlink to their Symptom Check that would be sent to those who were still active and within their 21-day symptom reporting window. Alert conditions were similar to the daily email alert, with the modification to begin the alerts the morning following enrollment.

We took the approach of providing both the email and SMS messages, but slight modifications could be made to provide an “either/or” approach. These would include modifying the Email/SMS alerts to only activate dependent on the SMS submission field. As alerts are our primary method of contacting participants, dual alerts for mail and SMS must be maintained.

Survey design modifications were made to the Symptom Check survey to optimize the display for smart phones: larger and more visible buttons were utilized to make data entry on mobile devices more convenient.

*Integrated Dashboard for Tracking; Reporting and Monitoring*

From within the REDCap application, there are a variety of ways to filter and sort data for analysis and export. The application includes a custom reporting feature which allows for highly specialized data filtering, but it does have limitations and often, much of the data analysis required to monitor compliance requires third party analytic suites or tools.

Methods were developed to provide a convenient and standardized process for identifying non-compliant participants, to document outreach efforts, and to communicate effectively with stakeholders on their progress. The tool identified as being the most familiar to the monitoring staff was MS Excel. A process was developed that would allow for data to be exported from REDCap via an Application Programmable Interface (API) directly into an MS Excel file. This file would then be used by monitoring staff to document daily phone outreach efforts.

Data was loaded into the Excel file via a Power Query accessing the REDCap API. The REDCap API is an interface that allows external applications to connect to REDCap remotely, and is used for programmatically retrieving or modifying data or settings within REDCap. A transformation of the data was preprogrammed that would produce a final dataset of non-compliant participants and their associated contact information. Call Center staff were trained on how to easily perform the extract, which required zero knowledge of coding, advanced excel knowledge, or authentication into REDCap. Monitoring staff were trained on how to document if a phone call was made to encourage participant participation and any notes that may assist in future efforts.

**Example API Query**

```
let
    redcapUrl = "https://REDCAPSERVER.org/redcap/api/",
    parameters = [token="REDACTEDTOKENSTRING",
    content="report",
    format="json",
    report_id="REPORTID",
    csvDelimiter="",
    rawOrLabel="raw",
    rawOrLabelHeaders="raw",
    exportCheckboxLabel="false",
    exportDataAccessGroups="true",
    returnFormat="json"],
    body = Text.ToBinary(Uri.BuildQueryString(parameters)),
    options = [Headers = [#"Content-type"="application/x-
www-form-urlencoded"], Content=body],
    Source = Json.Document(Web.Contents(redcapUrl,
options)),
```

```
        #"Converted to Table" = Table.FromList(Source,
    Splitter.SplitByNothing(), null, null, ExtraValues.Error)
    in
        #"Converted to Table"
```

Additional transformations to imported data were performed using the Excel Power Query tool. A report of all symptom report submissions was generated, then filtered to only include participants who had not completed a symptom report submission on the current day. Monitoring staff would generate this report at specific time each day, and use the associated staff contact information to generate a non-compliant participant list. Call center staff would then use this data to contact participants.

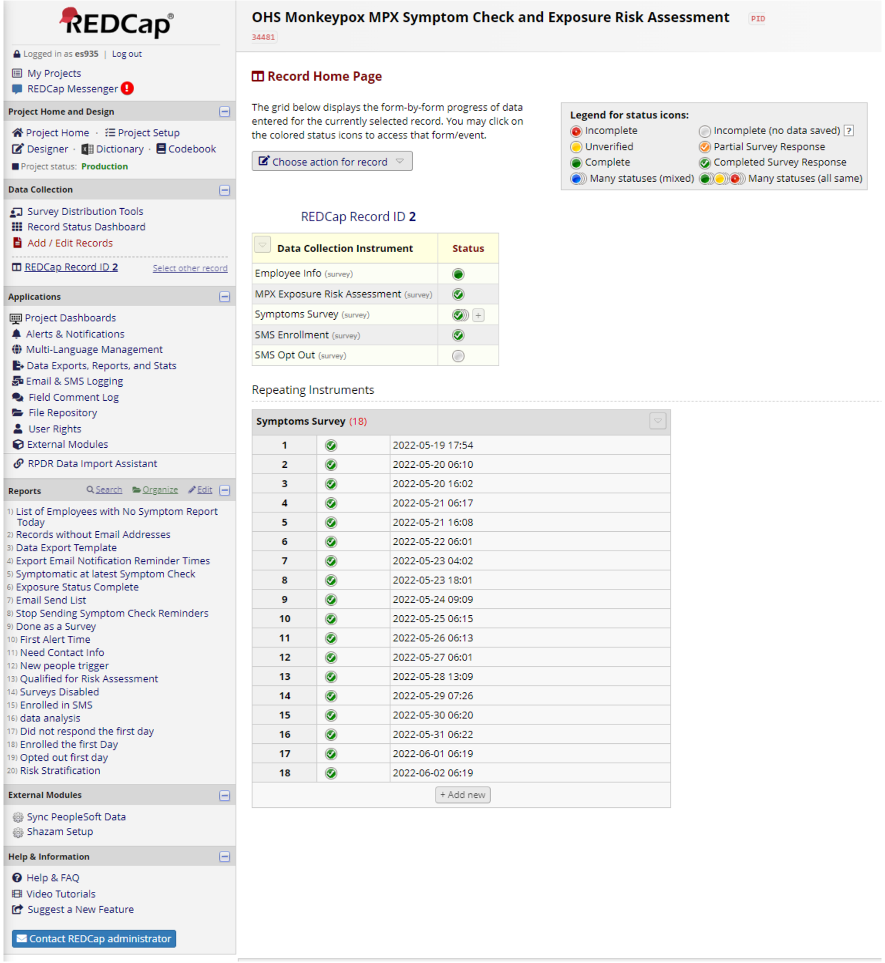


Additionally, many reports were created within REDCap that would allow stakeholders and technical staff the ability to monitor the actual symptom data that was being generated. Most notably a report was created that would return any participant who had reported being symptomatic at their latest check in. Options were discussed on how an email or SMS alert could be sent to monitoring staff to indicate a participant had reported being symptomatic but were deemed unnecessary as staff was actively monitoring the project.

*Miscellaneous items in the project*

Two items were implemented in this project for purely aesthetic reasons and are not vital to the functionality. The Shazam external module was activated and used on daily symptom surveys to modify the margins and spacing of items via CSS customization. A custom built external module, MGB survey theme, was implemented to apply MGB branding and formatting to all survey pages.

**Limitations**

An approach to only allow data entry during specific time windows each day was initially considered. This approach would require using the survey queue to detect the current time, and if to show the symptom survey. A bug was identified in how REDCap parses date values via PHP, preventing this capability. This bug was submitted to the consortium and acknowledged to be fixed in a future update.

Due to using alerts & notifications as the delivery method for daily surveys via the survey queue and limitations of repeating instruments, an automated survey reminder was unable to be set up. The REDCap consortium has announced that upcoming features to be added to repeating instruments may resolve this issue.

Implementation of the multi-lingual feature in REDCap was discussed but ultimately rejected as there was not a large need for the feature. Additionally, waiting for translations would have delayed project deployment and added complexity requiring further testing and maintenance.

**Summary**

The REDCap project build and any custom external modules code are available to other organizations to use and modify as needed. Customization may be indicated if an organization or entity is adapting local definitions of exposure or follow up based on responses. Required customizations will depend on the needs of an individual organization. At a minimum, an update of the alert & notification sender email address, wording custom to the organization on survey settings, and the authentication settings within the custom survey login external module will need to be made.

**Further Information**

Institutions and individuals who after reading the manuscript and supplement have additional questions can contact the MGB REDCap team at.
